# Clinician perspectives on implementing reduced preoperative fasting in Australia

**DOI:** 10.1101/2025.07.22.25331241

**Authors:** Oya Gumuskaya, Sahnoun Skendri, Nick Glenn, Rosemary Carroll, David Rowe, Jed Duff, Sarah Aitken, Amy Lawrence, Gerald Wong, Stefan Meisiek, Mitchell Sarkies

**Affiliations:** School of Health Sciences, Faculty of Medicine and Health, The University of Sydney, NSW, Australia; School of Nursing and Midwifery, Western Sydney University, Sydney, NSW, Australia; Honorary Lecturer, School of Nursing and Midwifery, College of Health, Medicine and Wellbeing, University of Newcastle, NSW Australia; Sydney Nursing School, Faculty of Medicine and Health, The University of Sydney, NSW, Australia; Clinical Nurse Consultant in Surgical Research, Surgical Services, John Hunter Hospital, HNELHD, New Lambton Heights, NSW, Australia; Clinical Director of Anaesthetics, Armidale Hospital, Armidale, NSW, Australia; Professor, QUT School of Nursing, Centre for Healthcare Transformation, QLD, Australia; Sydney Medical School, Faculty of Medicine and Health, The University of Sydney, NSW, Australia; Concord Institute of Academic Surgery, Concord Repatriation General Hospital, Sydney Local Health District, Concord West, NSW, Australia; Department of Anaesthetics, Royal Prince Alfred Hospital, Sydney Local Health District, Camperdown, NSW, Australia; Associate Professor, University of Sydney Business School, The University of Sydney, NSW, Australia; Implementation Science Academy, Sydney Health Partners, The University of Sydney

**Keywords:** Hydration, knowledge translation, safety, barriers, enablers

## Abstract

**Background:** Preoperative overnight fasting of patients (no oral intake from midnight until the time of surgery) is a potentially harmful practice; nevertheless, it remains common. Prolonged preoperative fasting is frequent, at times up to 24 hours of fluid and nutrition deprivation. International guidelines recommend reduced fasting time to improve patient outcomes, but this evidence is not well implemented. This study investigated clinician perspectives on two interventions designed to reduce preoperative fasting.

**Methods:** The qualitative study was informed by the Consolidated Framework for Implementation Research (CFIR). Semi-structured interviews were conducted virtually or in person, with a purposive sample of perioperative health professionals. Thematic analysis revealed codes, some of which were specific to the two reduced fasting interventions (SipTilSend and oral carbohydrate loading), organised according to the CFIR constructs.

**Results and Conclusions:** Twenty-one multidisciplinary clinicians were interviewed. Within the CFIR domains, adaptability enabled the tailoring of interventions to clinical contexts, while governance and policy updates supported adoption (Innovation). Progressive anaesthesia team leaders and leadership engagement drove change (Individuals). However, outdated policies and disincentives hindered progress (Outer Setting). Barriers included a lack of knowledge, while knowledge dissemination and clinician commitment to patient safety facilitated uptake (Inner setting). Champions among anaesthesia leaders and perioperative interdisciplinary collaboration played key roles in implementation success (Process).

University of Newcastle Human Research and Ethics Committee Approval No: H-2021-0328

## BACKGROUND

Excessive fasting of patients before surgery is a persistent issue in perioperative care, sustained by long-standing routines such as ‘nil by mouth’ (NBM) or overnight fasting (no oral intake after midnight) before surgery. These practices originated from Mendelson’s landmark 1946 study, which established the association between gastric aspiration during anaesthesia and maternal mortality[1]. Whilst routine perioperative fasting has improved aspects of patient safety, contemporary evidence recognises that prolonged fasting can also cause harm. Extended fluid and food restriction is associated with metabolic stress, insulin resistance, glycaemic imbalance, postoperative nausea and vomiting, anxiety and increased risk of complications [2–5]. Prolonged fluid restriction contributes to dehydration, hypotension, delirium, and delayed recovery, outcomes that disproportionately affect older and frail patients, prolonging hospital stays [6–9]. Multiple clinical audits have shown that fasting before surgery is often prolonged for up to 24 hours, placing vulnerable populations at clinical risk and delaying recovery [8, 10, 11].

Current guidelines recommend reducing preoperative fasting, allowing patients to consume solid food with protein and fat for up to six hours, and clear fluids up to two hours before surgery [12, 13]. However, translating these guidelines into practice is difficult due to the complex and unpredictable nature of perioperative care. Surgical schedules frequently and rapidly shift in response to unexpected emergencies, late case changes, or resource constraints. In this environment, perioperative staff may pre-emptively enforce extended fasting by keeping patients in nil-by-mouth in anticipation of theatre time [14, 15]. Additional barriers include staff shortages, inconsistent communication, and limited awareness of fasting protocols [14]. These factors contribute to the persistence of prolonged fasting despite guideline recommendations [2, 3, 16, 17].

To address these challenges, international protocols such as those developed by the Society of Enhanced Recovery After Surgery (ERAS) have been widely promoted [18, 19]. These protocols allow solid foods until six hours before surgery and recommend oral carbohydrate loading (OCL) to continue fluid and calorie intake until two hours before surgery. However, OCL has not been universally adopted due to limited evidence of patient benefit, cost concerns, and variation between individual practitioners and institutions [20, 21]. The fixed two-hour cessation window required for OCL is often impractical in real-world perioperative workflows, which limits its uptake and long-term feasibility [22].

Another emerging practice, referred to as “SipTilSend,” involves allowing patients to drink clear fluids, typically up to 200 ml per hour, until anaesthesia induction. This approach has gained interest both in Australia and internationally as a means to address prolonged preoperative fluid fasting [23, 24]. Early studies have reported reductions in postoperative nausea and vomiting, without an associated increase in aspiration or adverse outcomes, when adult patients were permitted to consume clear fluids until transfer to the operating theatre for surgery [24]. While promising, widespread adoption has been limited by the relative absence of high-level evidence on safety and effectiveness, and barriers to knowledge translation [5, 12, 25].

This study aimed to explore clinicians’ perspectives regarding implementing reduced preoperative fasting interventions and to develop strategies to inform future perioperative care practices.

## METHODS

This study used a theory-informed content analysis approach to explore clinician perspectives on the implementation of reduced preoperative fasting protocols [26]. Analysis was guided by the Consolidated Framework for Implementation Research (CFIR) outcomes [27, 28], with supplementary coding using the Capability, Opportunity, Motivation – Behaviour model (COM-B) to explore behavioural determinants within the CIFR Individuals domain [29]. The study is reported in accordance with the Consolidated Criteria for Reporting Qualitative Research (COREQ) checklist [30].

### Research team

The Principal Investigator (OG) conducted the interviews. The female interviewer has an MSc and PhD degree in surgical nursing, is experienced in qualitative studies, and was employed as an academic in a university during the study. While three out of 21 participants were acquainted with the interviewer via research collaboration, the study remained focused on their clinical experiences and perspectives on reduced fasting interventions. Importantly, the interviewer’s relationship with participants was not based on shared workplace or clinical area affiliations. The research team consisted of experienced clinicians and academics who are experts in their field and perioperative research.

### Participant selection and recruitment

A purposive and snowballing sampling strategy was employed to recruit perioperative clinicians from diverse professional backgrounds. Recruitment took place through professional association emails and newsletters, as well as through the authors’ personal social media and direct referral networks. Following an interim analysis of the initial sample, which primarily consisted of nursing participants, a second recruitment phase was undertaken. This phase targeted medical professionals, including anaesthetists, surgeons, and geriatricians, to ensure broader disciplinary representation and enhance the richness and depth of the data.

### Data Collection

Participants completed a semi-structured interview based on a CFIR-informed guide of 21 questions. This guide was developed and piloted by the research team to assess the factors influencing the implementation of reduced preoperative fasting practices [26, 27]. All participants provided informed consent. Interviews were conducted 1:1 via Zoom™ or in person between May 2022 and May 2024. Interviews were audio recorded, transcribed and deidentified for analysis. Basic participant characteristics were collected (participant gender, role, workplace and geographical location).

### Data Analysis

A directed content analysis approach was used, drawing on CFIR constructs. Abductive coding was also conducted to identify themes not captured by existing CFIR domains. CFIR consists of five domains and 39 constructs, providing a foundation for understanding implementation processes [27, 28]. Within the Individuals Domain, the COM-B model informed the interpretation of clinician behaviours related to capability, opportunity, and motivation [29].

Coding was performed using NVivo^TM^ software, with all transcripts independently coded by two researchers (NG and SS). Codes were mapped to CFIR domains, and additional codes were developed inductively as needed. Initial coding of two transcripts was used to establish agreement, with discrepancies discussed and resolved between coders, with oversight by OG and MS. The coding process also enabled the identification of innovation-specific constructs that did not align directly with the CFIR. After completing the coding process, the Principal Investigator (OG) consolidated the findings into comprehensive themes and subthemes.

### Ethics

Ethical approval was granted from the University of Newcastle’s Human Research Ethics Committee (Approval no. H-2021-0328). Participants were provided with a Participant Information and Consent Form, outlining the details of the study. All data were stored in a de-identified format to minimise reidentification.

## RESULTS

The study evaluated the barriers and enablers to minimising preoperative fasting in Australia. Twenty-one perioperative clinicians (six anaesthetists, 12 nurses, one surgeon and one orthogeriatrician) across Australia and an anaesthetist from the UK were interviewed (Table 1). The duration of the interview ranged between 20 to 57 minutes. None of the participants requested the transcripts to be returned for comment.

**Table 1.**
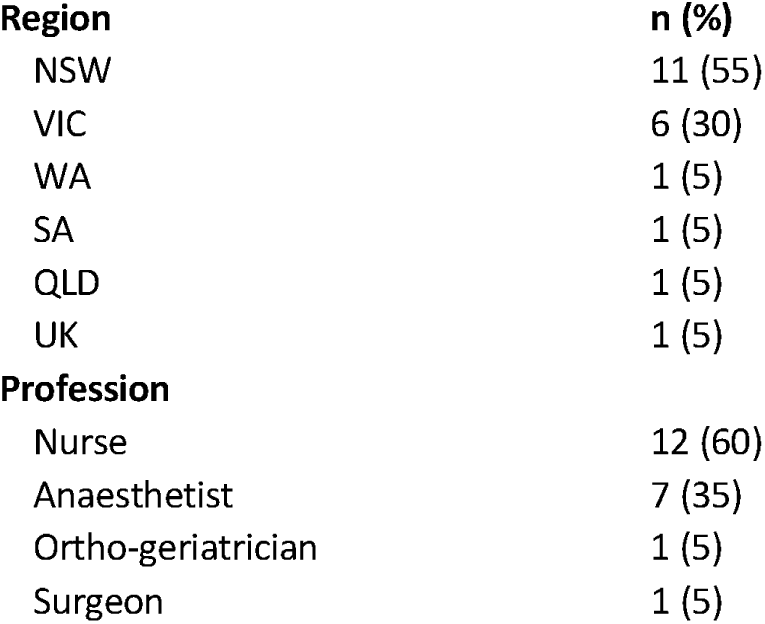
Participants demographics.

## INNOVATION DOMAIN

Figure 1 summarises constructs from The Innovation Domain. Participants identified credible sources like ERAS programs or the Australian and New Zealand College of Anaesthesia (ANZCA) guidelines as instrumental in promoting these innovations. While OCL was seen as evidence-based, SipTilSend lacked robust empirical data, relying more on anecdotal evidence and observational data due to the challenges of conducting large-scale trials. Despite this, both interventions were perceived to offer significant advantages over traditional fasting, including improved patient experiences, operational efficiency, and reduced healthcare costs.

**Figure 1.**
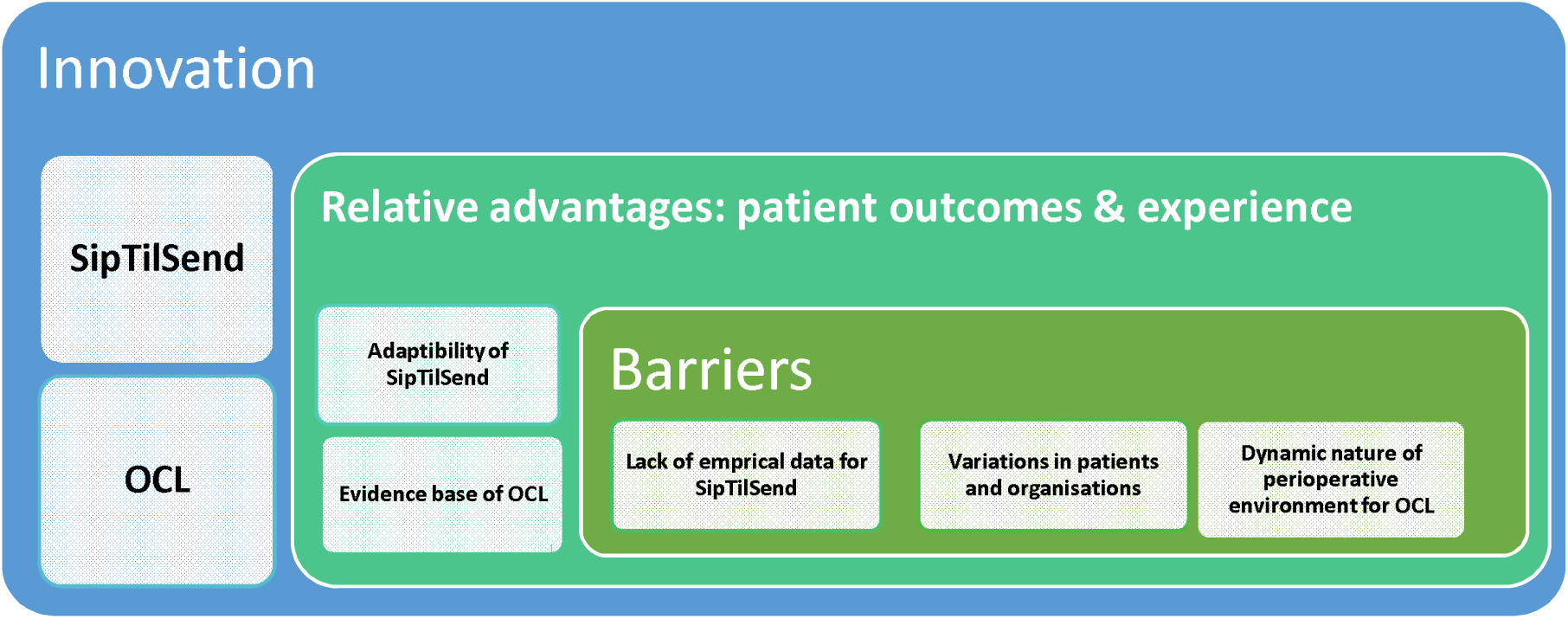
Innovation domain constructs emerged from analysis.

Several barriers were noted, particularly in implementing current fasting guidelines of two hours fluid fasting due to the dynamic nature of perioperative environments. SipTilSend was seen as more adaptable, allowing for customisation based on patient needs and clinical contexts. Its trialability was also a strength, enabling small-scale testing before broader adoption. The complexity of implementation posed challenges due to patient variability, institutional differences, and integration with existing protocols. Clear communication, simple instructional materials, and cost considerations were highlighted as essential for the successful adoption and sustainability of these innovations. Participant statements corresponding to each construct are demonstrated in Table 2.

**Table 2.**
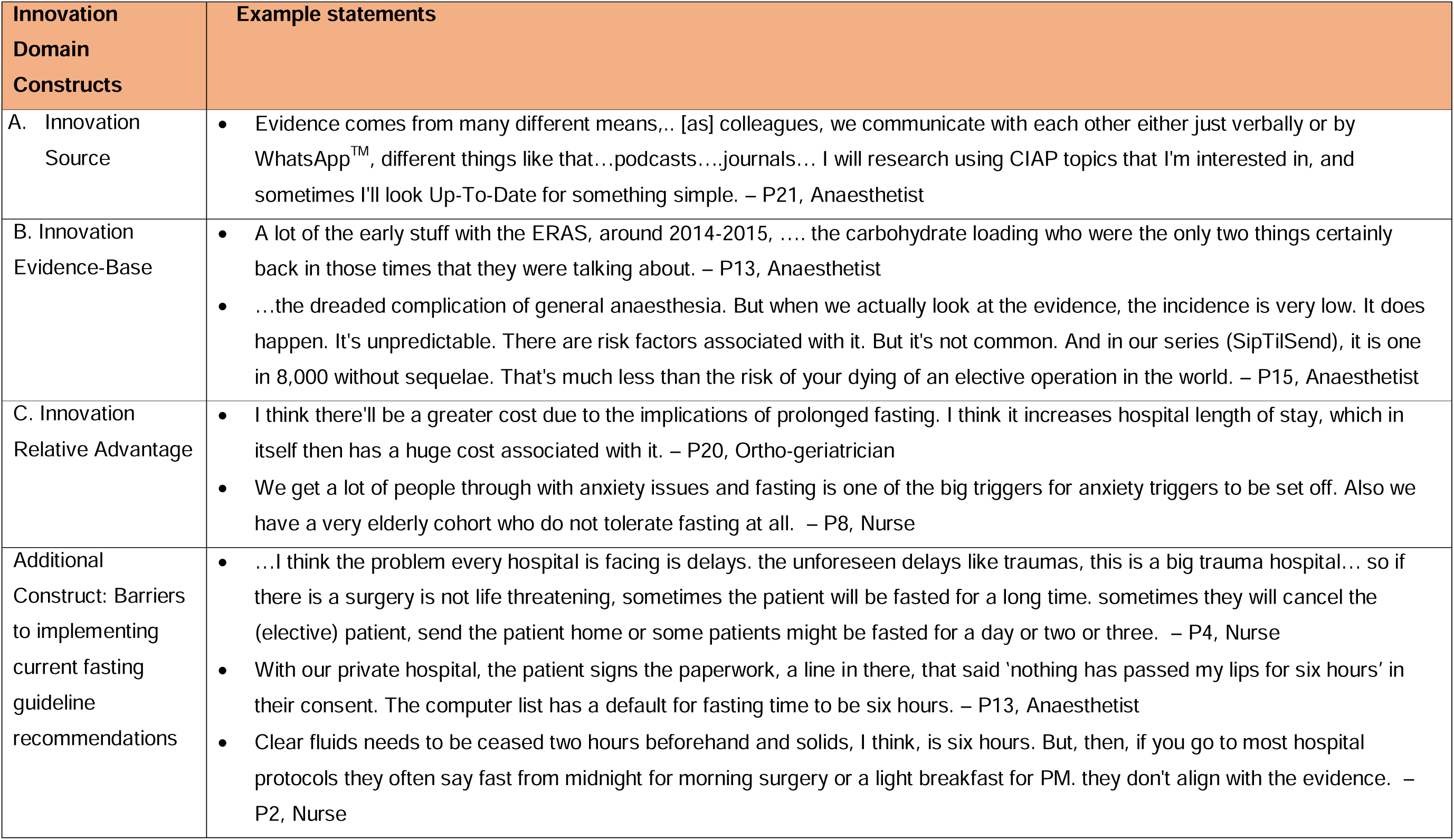
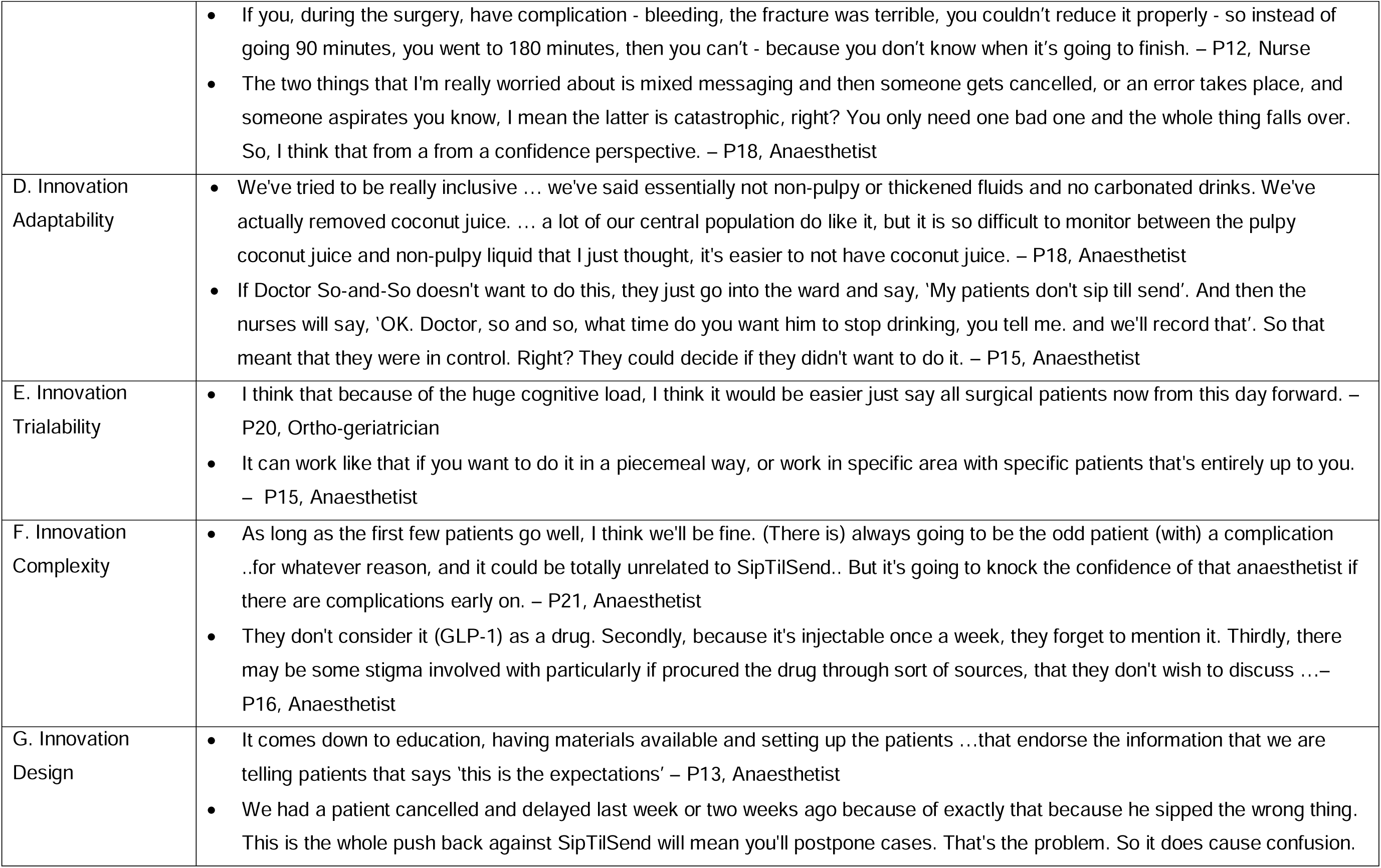
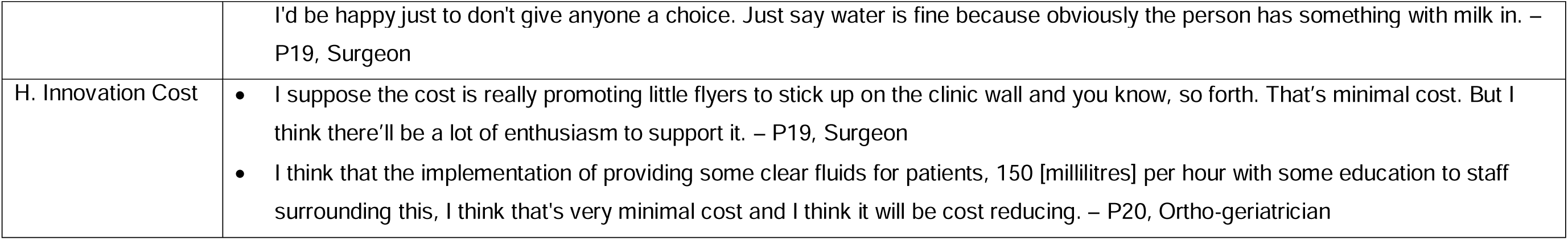
Innovation domain constructs and example statements.

## II. OUTER SETTING DOMAIN

**Figure 2.**
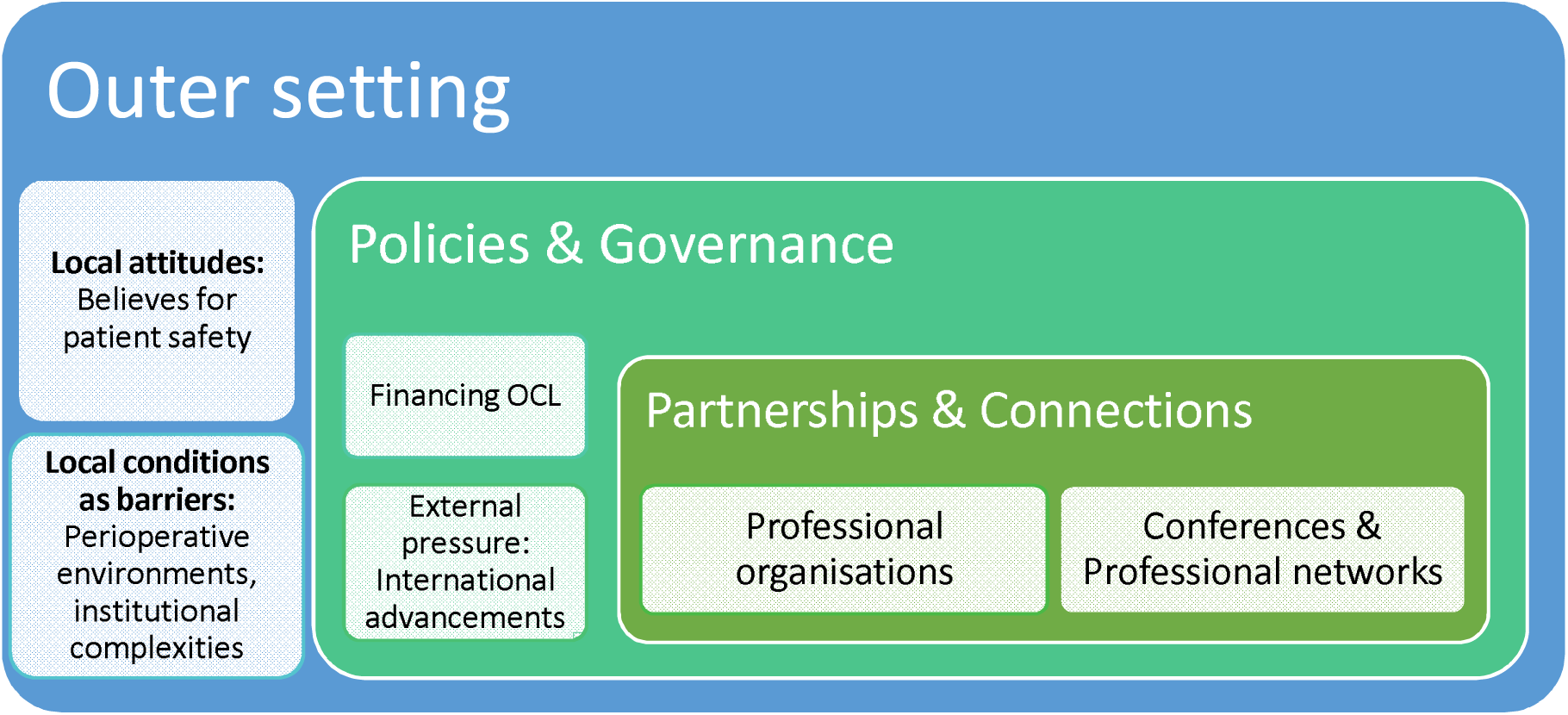
Outer setting domain constructs emerged from analysis.

Economic and institutional complexities within perioperative care environments posed significant barriers, particularly for interventions like OCL, necessitating adaptive strategies to navigate these challenges.

External partnerships and professional networks played a crucial role in enabling knowledge exchange and fostering a unified approach; however, policy and governance structures presented barriers. Participants highlighted the need for a local policy update, aligned with professional guidelines. The outdated procedures and complex governance processes hindered the adoption of innovations like SipTilSend.

Financial considerations also influenced implementation, with funding required for OCL drinks and potential adverse event management under SipTilSend.

International trends and external factors, particularly those from the UK and European ERAS guidelines, have significantly influenced local practices. These trends helped shape clinicians’ understanding and created performance pressures tied to operational efficiency and service goals. Demonstrating the cost-effectiveness of SipTilSend, such as reducing surgery delays or cancellations, was seen as essential for its broader integration into clinical practice. Participant statements corresponding to each construct are demonstrated in Table 3.

**Table 3.**
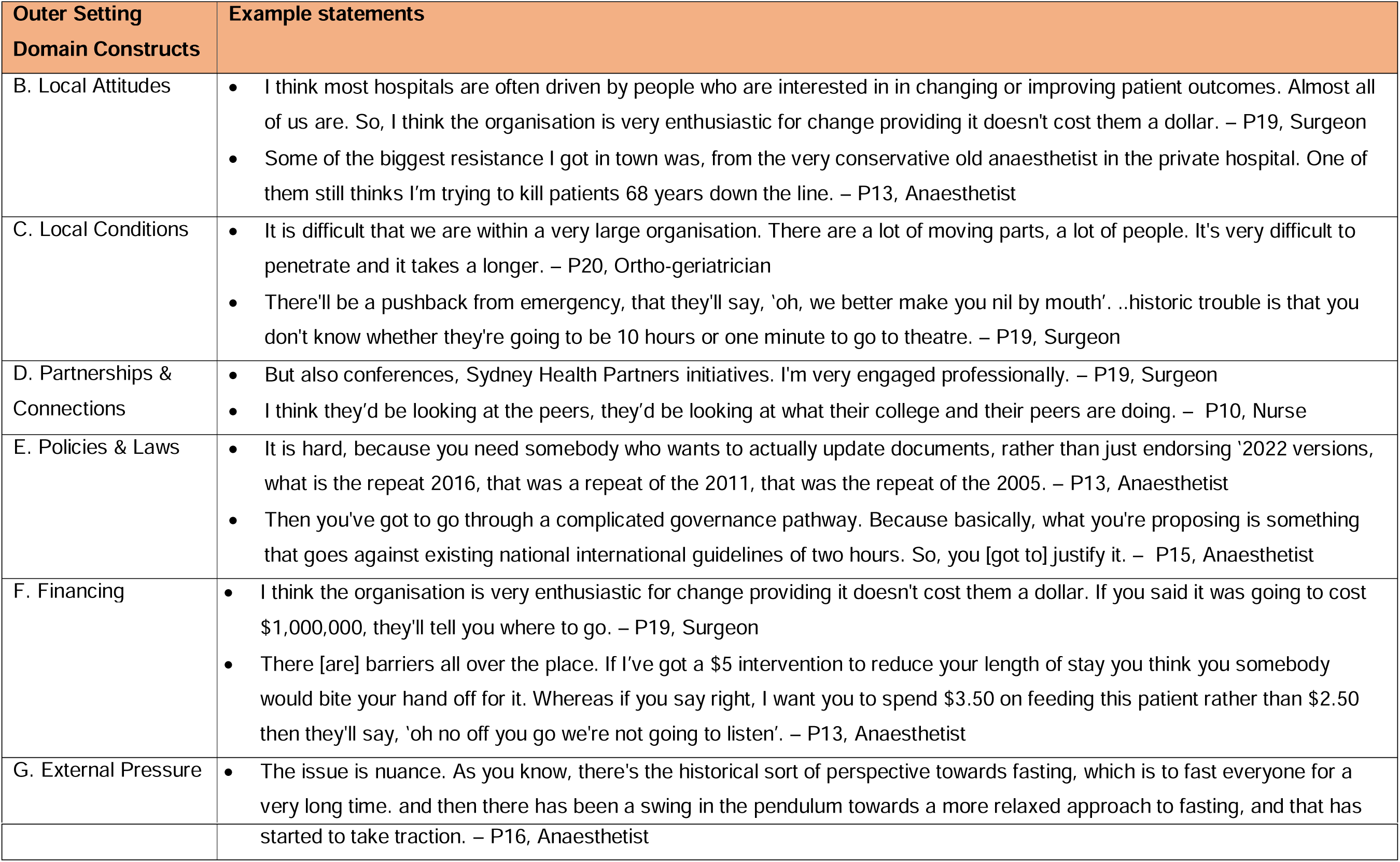
Outer setting domain constructs and example statements.

## III. INNER SETTING DOMAIN

**Figure 3.**
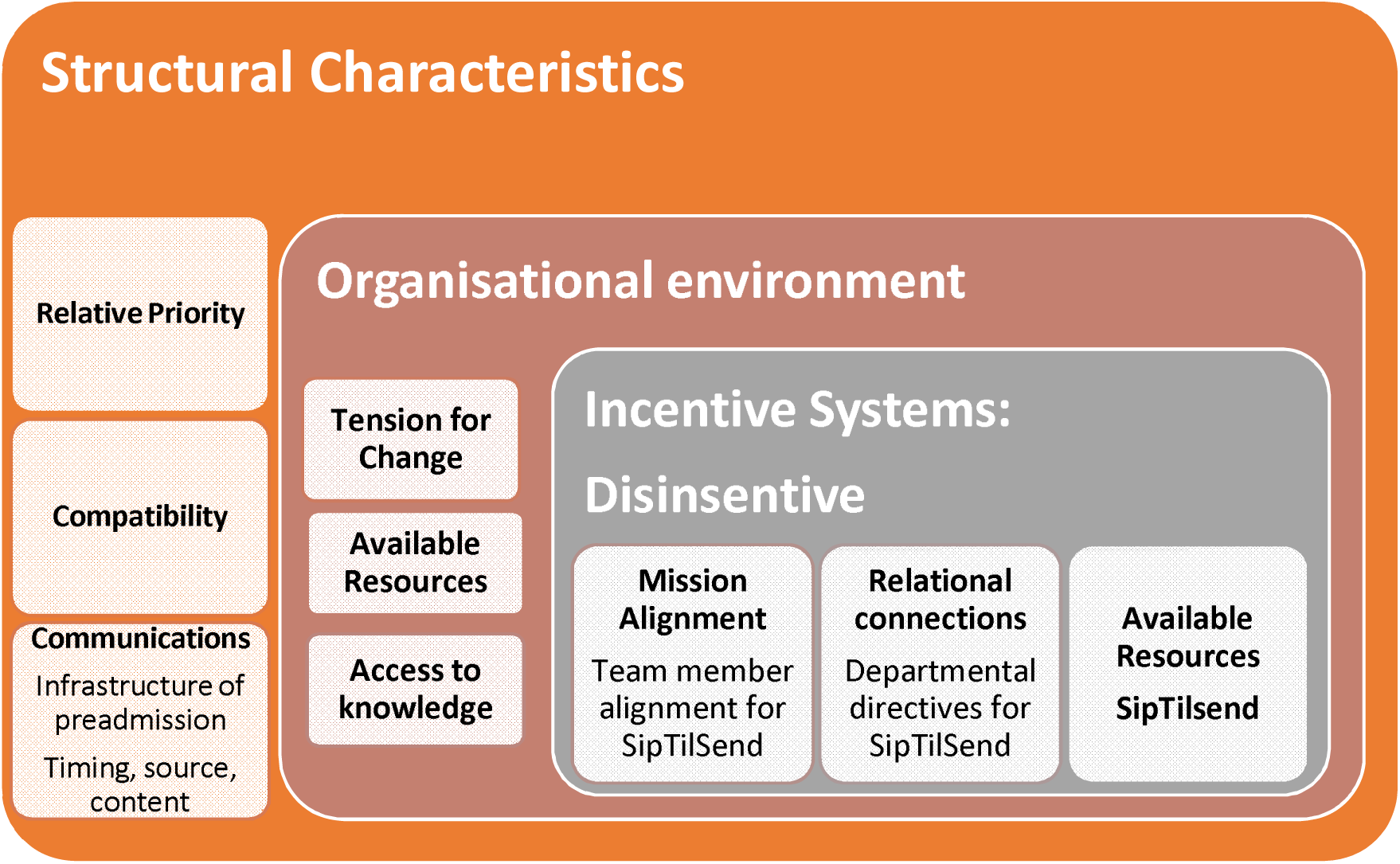
Inner setting domain constructs emerged from analysis.

The effective infrastructure of preadmission clinics and flexibility in surgical preparation were required for the implementation of SipTilSend. The importance of clear roles and responsibilities, especially in managing emergency cases was noted. The timing, source and content of preoperative communication with patients varied among patient cohorts. How, when, where, and by whom these instructions were provided varied significantly among cohorts, theatres or institutions, such as before or on patients’ arrival at a hospital for surgery. Available information technology for preoperative patient communication also appeared to be a significant structural factor (Communications).

The quality of formal and informal relationships was critical for the SipTilSend implementation process. Participants noted the need for departmental directives to mitigate resistance and foster collaboration. It was stated that establishing connections within the institution and allowing practitioners to voice concerns and preferences on SipTilSend or OCL was beneficial. On the other hand, the hierarchy between clinicians hindering communication, coupled with the high staff turnover, can present challenges. Unified departmental directives were a method to reduce the anxiety to support the implementation of SipTilSend (Relational connections, Incentive systems). Communication within and across settings was an essential part of any change in practice. Participants identified multiple channels, such as group messaging applications on personal devices, podcasts, or written materials, as tools for disseminating information for clarifying reduced fasting instructions, and clarification of intervention steps was desired for both clinicians and patients. Clinicians raised concerns about patient miscommunications regarding instruction for fasting, which was a common occurrence even with printed material. Team meetings, though not preferred, were suggested to enhance engagement with implementation.

Participants indicated their organisations supported new approaches, though some resistance to change was also observed. Participants emphasised the importance of aligning SipTilSend with individual patient needs, reducing risk while maintaining flexibility in implementation practices. Medical professional participants mostly felt valued and respected within their workplace environment. Most workplaces were open to implementing new approaches, though there were various responses, with some nurses indicating a lack of support for change in practice (Organisational environment).

Current fasting practices were often more conservative than evidence based recommendations, and failure to implement such strategies resulted in prolonged fasting causing distress to patients. While the desire to advance the preoperative fasting practice was expressed, challenges to start discussions among the key stakeholders, such as registrar medical officers was also indicated.

It was suggested that SipTilSend may not be suitable for all patients, that the risk-benefit ratio be carefully considered for specific groups, including obstetric patients, emergency cases, patients with reflux, and patients taking GLP-1 receptor antagonist medications.

Interviews stated that the busy nature of the clinical environments results in the planning and implementation of new practices being of low priority. Some participants indicated that the increased operating room efficiency due to reduced cancellations would contribute to the desire to implement SipTilSend. The environmental stresses were described as significant barriers to OCL uptake, which may result in clinicians reverting to old routines to reduce their cognitive load. One interviewee discussed a disincentive: how nursing staff may not provide patients with fluids within SipTilSend or OCL, out of fear of being reprimanded. This cognitive dissonance compromised patient care through prolonged fluid fasting. Other participants noted there was no incentive within their hospital; however, if there was a strong champion, their efforts may be recognised(Insentives).

Interviewees expressed a significant need to address oppositions with key stakeholders. Challenges did present, e.g. the presence of outdated opinions within the team and the high rotation of junior medical officers, necessitating the need for re-education to address practices learnt in other teaching hospitals. In seeking alignment, the necessity of ongoing provision of information was indicated, including the motives and rationale behind the change. Interviewees expressed the need to bring all members of the multidisciplinary team on board to ensure alignment to promote better patient outcomes during the intervention.

Several sources were identified for accessing knowledge, monthly meetings, newsletters, journal clubs and emails, all of which can be sporadic. In many of these sources, clinicians’ involvement may depend on their interest. Medical practitioners were kept updated on evidence through group messaging applications, podcasts, journals and professional websites. Patients access information on the wards, through staff and printed material. Posters were noted as effective for clinicians of changes in fasting practices. Participant statements corresponding each construct were demonstrated in Table 4.

**Table 4.** Inner setting domain constructs and example statements.

| III. INNER SETTING<br>DOMAIN | Example statements |
| --- | --- |
| A. Structural<br>Characteristics | • Most tertiary hospitals in Sydney, the area that they (patients) arrive at in the hospital for elective surgery is called the TPU. – P18, Anaesthetist<br>• Paediatrics ward will call the theatre and ask up until what time they can have clear fluids. There's no protocol for adults. Sometimes if a patient ask for a drink, they might call theatre. – P14, Anaesthetist |
| B. Relational<br>Connections | • The trouble is we have such turnover of nursing staff as well, near now so many junior RN's stuff on the ward, it's a challenge on that front. – P13, Anaesthetist<br>• We are an academic institution, need to facilitate research..., so I'm not really giving people the option of saying, 'I don't want to participate'. This is now a departmental directive. I also had to consider people's opinions, and there is some anxiety. Only a week ago I had anaesthetist who cancelled surgery because a patient had had clear fluids within two hours, and people got pushed to the next day. –P18, Anaesthetist |
| C. Communications | • In terms of size, that just takes time to penetrate and it's multi-pronged. So, merch! You know it's in services. It's word of mouth. It's just it's just time and it's patience. Persistence. –P20, Ortho-geriatrician<br>• There needs to be a better line of communication between what's happening in theatres and someone on the ward who can then manage the fasting status. – P16, Anaesthetist<br>• I actually think those weekly meetings have really been good because it's always now in the in front of my mind. I see you every week. I see you on my screen. We talk about it. We talk about some of the barriers, the limitations and things have gone well. Some things have not got so well, and I think that's kept it in the forefront of my mind. – P20, Ortho-geriatrician<br>• We have staggered admissions during the day. We don't tell patients the time in preadmission clinic or when they're coming in. They ring the hospital beforehand. Day surgery have the running order of the list. Day surgery give the patient two times, one which is a time to come into the hospital, and the other is a time that they can keep drinking up |
|  | until. – P13, Anaesthetist |
| D. Organisational Environment (Culture) | • The motivation to do anything is just because you think it's the right thing to do. - P20, Ortho-geriatrician<br>• I'm certainly respected by my head of department by my peers. Yeah, I think so, yes. - P21, Anaesthetist<br>• I feel heard, yeah. – P20, Ortho-geriatrician |
| 1. Deliverer-Centredness | • You know there'll always be somebody. Though we say that's fine, you know. The other area that we've used the preop drinks is in endoscopy and bowel prep. So rather than broth and jellies and things like that, we tell the patients, four bottles of preop the night before once you've started your bowel prep and then two on the morning of surgery. And that ...means that with our diabetic patients.. don't stop their medications. – P13, Anaesthetist |
| 2. Learning-Centredness | • People sort of said 'We've heard this lots of times before'. And I said, Listen, I said, 'Look, guys, the only way to change this is not to reduce it to 1 h, or half an hour, or 10 min. It is to go SipTilSend, and this means the patient can drink, sip water right until their sent to theatre and I and I explained the physiology, and to my surprise, the majority of the people on the call agreed. They could see the sense - P15, Anaesthetist<br>• ...anaesthetic department, we have weekly nursing meetings....Facilitator: All team members? Interviewee: Yes. Monday morning at 8:00, a lot of it's education on that day but there is a chance for the managers to give a bit of staff meeting as well. – P9, Nurse |
| E. Tension for Change | • I think there's some interest to implement the change ... Yeah, there's strong, proactive people within the department. – P21, Anaesthetist<br>• instead of getting done at eight o'clock, they'll be getting done at 12 o'clock. Because you have less theatres, less staff, but then we try to do the same number of patients. Especially Q1 surgery. They come already from far away, they already in the hospital [unclear] get them done. So, it is getting worse. Like the waiting time is a bit longer at the moment because short of staff, I think. – P12, Nurse |
| F. Compatibility | • they are not quite the same as the general population in terms of fasting and gastric emptying. So we traditionally describe the obstetric population as never truly fasted. – P18, Anaesthetist |
|  | • But [you got to] be a bit selective with some of the patients and patients with no gastric delay problems people who are diabetics you know. So obviously, people would get outward obstruction. But now, as I mentioned a lot more people now, are actually on the glutides, and they won't necessarily say that they're on them. – P16, Anaesthetist |
| G. Relative Priority | • Particularly the trauma list, or for orthopaedics, is pretty bad. You have patients fasted for three or four days in a row all day. It is always difficult when there's a list like that that the priorities change all the time, but I do think they could probably make decisions earlier in the day about who they're definitely not going to do, and certainly with clear fluids like as an excuse for that. – P14, Anaesthetist<br>• if you could target how you manage those [diabetic] patients, how you improve say their calorific intake, their sugar control with the millions of people who are Type 2 diabetic around the world, that will be a massive thing to target. – P19, Surgeon |
| H. Incentive Systems (Disinsentive) | • Nurses are petrified on the ward. Every nurse will remember the one patient that they gave a drink to, and they lost their slot of the emergency board. And they are never going to repeat that mistake in in their life, because one anaesthetist getting up there and yelling and carrying on because they had a drink. It will mean that every other patients they will look after for the next decade will have their care compromised. – P13, Anaesthetist<br>• Recognitions or rewards? I don't think so. Well, you know, the hospital does have different things. Whether somebody would be recognised for it. I can't say. Yeah. So if there was a really strong champion, yes, they might get recognised for it within the hospital. – P21, Anaesthetist |
| I. Mission Alignment | • So that's your starting point. You've got to try. And you've got to achieve a consensus within the anaesthetic department, and you've got to explain where you're coming from and what the reasons are, and all that stuff because unless you've got that, you're really going to struggle... So that's step 1. – P15, Anaesthetist<br>• ...which I think would be beneficial for all our patients if they instead of fasting for eight hours, 10 hours, 12 hours, if they can just have a sugary drink or a cup of tea with honey, three hours before their surgery as opposed to waiting four or five hours, I think change could be implemented. But we'd have to definitely sit down and have a chat with our |
|  | anaesthetists. – P1, Nurse |
| J. Available Resources |  |
| Funding | From the anaesthetic side, who would take that on board as part of their role? Because other hospitals have worked at a structure differently, like the way that a lot of the consultants are ... only contracted procession. I find in general, there's a lot of reluctance to put any effort into that kind of beneficial thing for the whole department, because people [are] not paid for it. – P14, Anaesthetist |
| Materials & Equipment | it certainly be quite easy for the clinic nursing staff and anaesthetic staff to give the patient some information. Flyers would definitely be good, because I think a lot of it would just be that education for staff. – P14, Anaesthetist<br>It's pretty much given that there will be some ultrasound machines, but there always not enough. I've never seen a department of anaesthesia that is so well resourced that here's there's always fighting for the ultrasound like, who's got it last – P16, Anaesthetist |
| K. Access to Knowledge & Information | Everywhere, I mean every everywhere. But I mean, I suppose I'm heavily involved in in teaching and training at both the medical student and the more postgraduate level. So, I guess I'm learning all the time and also passing that information on so. But also, conferences. – P19, Surgeon<br>We also have a weekly education meeting in my department. I also do a few journals email me whenever they release their new addition, so get a kind of a monthly report. These are the new articles that we're publishing this month. So those are the main sources, I think. – P17, Anaesthetist<br>So evidence comes from many different means, obviously we communicate with each other either just verbally or by WhatsApp™ .... I listen to a lot of podcasts. I read, journals, and then I will research using CIAP topics that I'm interested in, and sometimes I'll look Up-To-Date for something simple. – P21, Anaesthetist |

## IV. INDIVIDUALS

**Figure 4.**
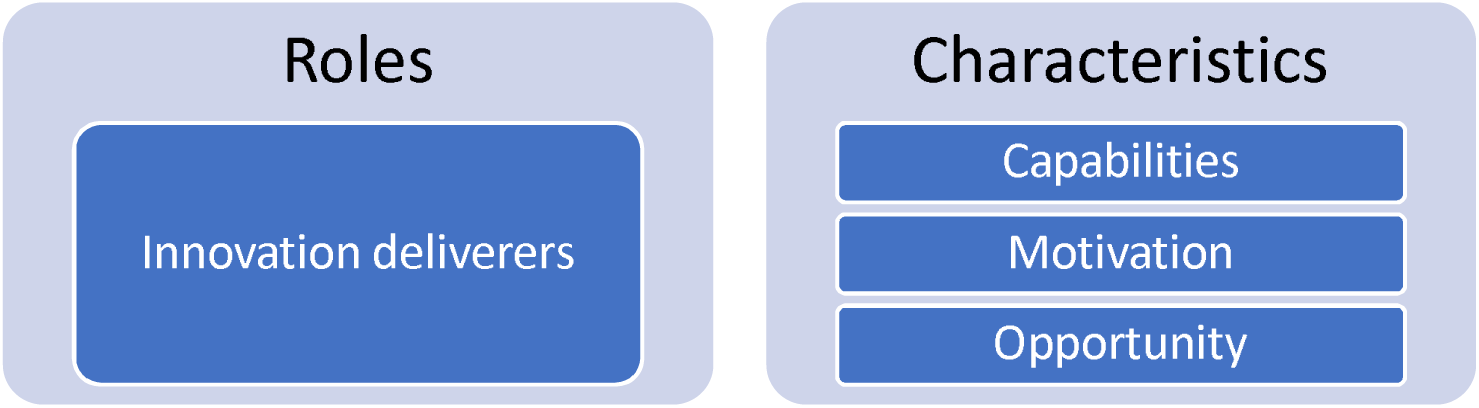
Individuals domain constructs emerged from analysis.

In elective surgeries, the Day Surgery Unit nurses were identified as key stakeholders, as they communicate new care approaches to patients and other healthcare professionals (Innovation deliverers). Their understanding and support are recognised as vital in the implementation of reduced preoperative fasting interventions. Ward nurses were also identified as key communicators within perioperative settings to facilitate the staggered implementation of STS across surgical units and departments. Patients undergoing surgery are the recipients of STS or OCL, and their concerns or suggestions should also be considered at the planning stages of implementing reduced preoperative fasting.

Support from department heads and staff specialists was identified as crucial. The need for strong leadership and evidence advocacy was emphasised, described as ‘strong people’ within the department. Leaders engaging with departments and presenting ideas and evidence (including retrospective local data) appeared crucial for gaining support. Perianaesthesia team members, including anaesthetists and surgeons, nurses, nurse educators, and administrative staff involved in developing the fasting protocols and executing the interventions, were expressed as pivotal. However, resistance from conservative anaesthetists was noted as a barrier. Their concerns about potential harm from SipTilSend highlighted the need for targeted engagement strategies. Clinical facilitators, such as preceptors and educators, through professional development initiatives, contribute to implementation success through mentorship, audits, and staff training. Allied health, nursing and other health professionals were all considered important implementation leads for the innovation. As such, there is a need to be open to the team’s input during in-services and education sessions. This provides an opportunity to discuss the effectiveness of the intervention, its safety and the logistics associated with implementation. Participant statements corresponding to each construct are demonstrated in Table 5.

**Table 5.** Individuals domain constructs and example statements.

| INDIVIDUALS<br>DOMAIN | Example statements |
| --- | --- |
| Innovation<br>Deliverers | • Day surgery nurses the day before. They will ring them up and check and tell them what time to come in and what time to fast from because when we see them in the clinic, we don't know the order of the list. So the patient may be first on the list, they may be last on the list or somewhere in between. – P21, Anaesthetist<br>• Our patients are actually quite compliant. If you give them written instructions and tell them what to do, they'll follow them and actually it was the elective patients coming into hospital admitted, on the day of surgery was easy one because we've got just a little fasting brochure that says, 'go out and get this drink'. – P13, Anaesthetist |
| **Characteristics Subdomain** |  |
| Capability | • There has been another one of the nurse educators from theatres redid some other posters for us about two years ago and they are still up and around in the place. – P13, Anaesthetist<br>• Some of the bosses do quality improvement projects or audits. I don't think there is a specific requirement for consultants. – P14, Anaesthetist<br>• I think it's an upskilling of the anaesthetist. So, when patients coming to the anaesthetic bay, then you can do a quick gastric ultrasound to actually determine whether the patient is actually fasted enough. And that will be a solution to whether patients have been fasting long enough. – P16, Anaesthetist |
| Motivation | • Some of the biggest resistance I got in town was, from the very conservative old anaesthetist in the private hospital. One of them still thinks I'm trying to kill patients 68 years down the line. – P13, Anaesthetist<br>• it's about an anaesthetist having a backseat and not putting our message front and forward and telling patients actually ... with the fasting that's not the most convenient for the running of the list, but the fasting that achieves the best outcome for the patient. – P13, Anaesthetist |
| Opportunity | • So, I'm in charge of the JMO education for geriatric ... I would implement an in service essentially and an education session for the |
|  | JMOs. So that's how I do that. And then there would be also that would be my input and then I would encourage in services for the nurses, allied health, all stakeholders essentially. – P20, Ortho-geriatrician<br>• ...we'll see them (patients) about their three or four weeks before they're coming into theatre for the day, and we'll sit down and talk about the anaesthetic, and we'll give them the instructions. We've got about a two-week lead time which is around this all the time, we could be useful for giving them. – P13, Anaesthetist |

## V. IMPLEMENTATION PROCESS

**Figure 5.**
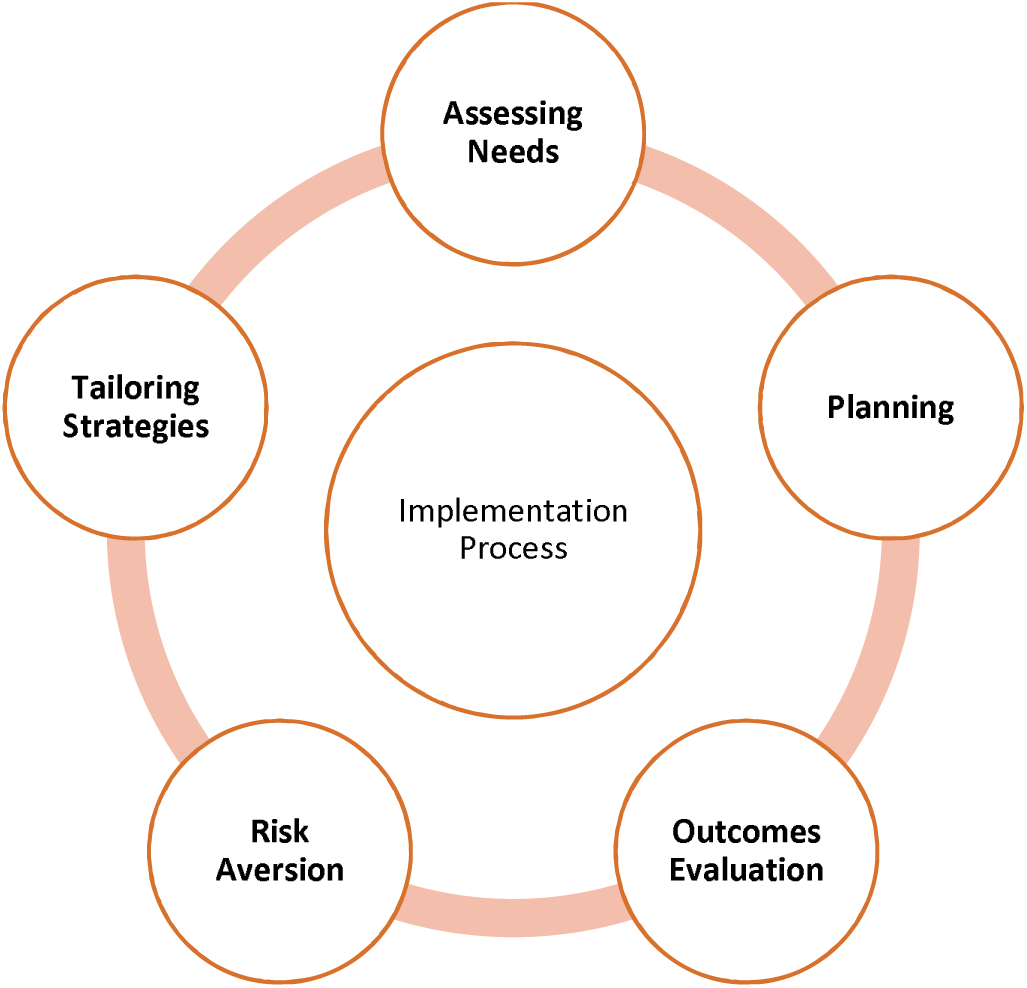
Implementation process domain constructs emerged from analysis.

Audits and surveys were found useful in gauging views and assessing needs before and during the implementation process. This included collecting data on patient’s hunger, thirst and nausea as a way of understanding current patient satisfaction. An audit conducted by the department was ‘fairly consistent’ in support of SipTilSend, re-occurring every twelve months; a survey to gauge the willingness and views of clinicians (especially anaesthetists) before and after implementation. This suggests that collecting quantitative and qualitative data through audits, surveys, etc., should be continuous to assess the needs of both deliverers and recipients.

Various approaches to plan implementing an intervention, such as SipTilSend, presented advantages and disadvantages. When deciding between a staggered departmental approach versus a hospital-wide implementation of SipTilSend, participants considered that launching SipTilSend requires a minimum of six months of planning. A staggered rollout beginning with a single department, while achieved in a shorter period, may create confusion among patient groups excluded from the initial phase, especially when adaptations of SipTilSend differ across departments. Concerns were raised about whether allowing multiple types of drinks might lead to confusion. Participants also discussed the need to draft clear guidelines, acknowledging that processes such as peer review could take several months. Additionally, they emphasised the importance of planning for emergency cases and considered the structural characteristics of each department, including the high number of clinicians with diverse concerns and experiences, which could influence the success of implementation.

In the process of implementing an innovation aimed at minimising fluid fasting, some interviewees stated that a top-down organisational approach may not be the most effective at promoting change within an organisation. Allowing wards to create tailored individualised strategies around aligning team members, promoting individual clinical decision making and allowing for patients to opt out drives participation in change and builds ground-level support for the innovation during implementation. When tailoring strategies, consideration should be given to the fact that each clinician has their own individual preference.

Example statements pointed to the necessity for patient safety and satisfaction evidence. While participants acknowledged that quantitative data may be needed, looking at aspiration could be futile due to a very low incidence rate requiring very large samples to demonstrate safety. It was stated that collection of observational data may be the only feasible option. It was noted that there will always be patients experiencing complications, and these may impact the confidence of anaesthetists if these complications coincidentally occur in the early stages of the SipTilSend implementation.

Participant statements corresponding each construct were demonstrated in Table 6.

**Table 6.** Implementation process domain constructs and example statements.

| IMPLEMENTATION<br>PROCESS DOMAIN | Example statements |
| --- | --- |
| B. Assessing Needs | • We did an audit, we sort of just went on the bandwagon of 'you've got a month to carry on starving the patients as you normally do'. We just threw together a real quick list of 'how your patients feel as they're arriving' in data. So we asked them about hunger and thirst and nausea and stuff like that. – P13, Anaesthetist<br>• Most things go via our head of department. She's usually aware of any policies and protocols, and certainly could find out the information easily. – P14, Anaesthetist<br>• These are all anaesthetists, right? So immediately, like the week after, we said 'Right, we're going to survey you, and we're going to ask you two simple questions. Would you be happy for your patient to sip water, one glass of water per hour until they go to theatre. Yes or no. Second question, 'would you be happy for them to have a hot drink, a cup of tea with 15 mL of milk in it when they arrive in the hospital. Ninety percent said yes to both of them. – P15, Anaesthetist |
| D. Planning/Engaging | • First thing we've [got to] do here is ..... go through a departmental meeting if you're [going to] make a change like SipTilSend because it's meaning that it's not just my patients who are going to be impacted if it is just my patients, it's really messy. – P21, Anaesthetist<br>• We debated long and hard about ... how to introduce it. whether we should do it in a specific group of patients, a specific ward, a specific hospital. And there are pros and cons for that. But we spoke to the nursing staff on the ground and the managers, and they said, 'we think you should go all on one day as long as we put the work in beforehand to make sure that everyone is educated'. – P15, Anaesthetist |
| E. Tailoring Strategies | • You need to have ground level support and believe that this is a viable option. And you also need to have a top-down approach as well, saying that these guidelines that people should follow and that will help to drive participation change as well. – P16, Anaesthetist<br>• The other piece of advice I'd give you is, don't try and do top-down organisation. So we didn't tell every single ward how to do |
|  | it. And so, rather than telling them how to do that, we said, ' think about how; ..what would work for your board, your department, and you do it your way'. So that gave them some kind of involvement in the whole thing, and it became easier that way. – P15, Anaesthetist<br>• You know there'll always be somebody. Though we say that's fine, you know. If Doctor So-and-So doesn't want to do this, they just go into the ward and say, 'My patients don't sip till send'. And then the nurses will say, 'OK. Doctor, so and so, what time do you want him to stop drinking, you tell me. and we'll record that'. So that meant that they were in control. Right? They could decide if they didn't want to do it. – P15, Anaesthetist |

## DISCUSSION

The analysis identified key enablers and potential barriers associated with implementing reduced preoperative fasting protocols. During the study period, there was a shift in how reduced preoperative fasting was viewed. Some participants from several sites refocusing their efforts towards the liberal fluid fasting model (SipTilSend), whilst others were aiming to reduce preoperative fluid fasting to two-hours with oral carbohydrate loading (OCL). The coding framework reflected this shift in intervention scope.

Several factors constrained the acceptance and sustainability of reduced fasting practices. For the SipTilSend model, the lower level of supporting evidence, misalignment between new protocols and existing hospital policies, and the need for updated guidelines from key professional societies (like the Australian and New Zealand College of Anaesthetists) were identified as barriers. The identified main barriers to the two-hour clear fluids policy or preoperative OCL recommendations related to structural feasibility, such as unpredictable theatre timing, the lack of resources for providing preoperative patient education and fasting instructions, clinician workloads, governance concerns and management of daily surgery lists. For all models of reduced fasting, enablers included visionary peri-anaesthesia team leaders, an interest in improving patient safety and experience, continuous dissemination of information to stakeholders, and perioperative team collaboration.

Despite current recommendations or the SipTilSend initiative, prolonged preoperative fasting is a prevalent issue, and the literature has not been conclusive for strategies to support translating knowledge [22, 31–35]. Society of ERAS guidelines, which promote OCL and reduced fasting, have been rapidly adopted across the globe; however, there are significant variations in practice, and the evaluation of the barriers to uniform adoption remains unresolved [18, 19].

Previous studies, though heterogeneous, inform us that a main issue regarding the barriers to translating knowledge into practice is the logistics within the complex perioperative hospital systems. The unpredictability of the theatre times and ever-changing operating lists were upcoming themes in the literature, which was supported by the results of this study [22, 36, 37]. To overcome these barriers, considering the de-implementation of unnecessary fasting practices, SipTilSend has been increasingly adopted [24]. However, key challenges in adopting these innovations included the lower level of evidence for SipTilSend, conflicts with existing policies and guidelines, and the need for professional society support.

While the advantages of SipTilSend over the current rule of overnight nutrient and fluid deprivation before an operation seem clear to many, the uptake of this approach is slowed due to leadership, communication and organisational factors. In terms of leadership, managers might hinder change by underestimating the work required to change a practice, overestimating the organisation’s capacity to change, and misjudging how stakeholders perceive the change[38]. While the research results call for leadership from perianaesthesia, this could be facilitated through choosing the best implementation approach in terms of local or systemic rollout and pace. Furthermore, leadership could consist of encouraging, piloting and experimenting to build local knowledge, assimilation and integration that overcomes a ‘not-invented-here’ resistance. Leadership means balancing stretching and stress to keep change at a pace that the organisation can take.

A consistent concern across the literature, echoed by participants in our study, is the lack of awareness and understanding of institutional and national fasting policies [21, 38]. Several participants indicated that neither they nor their colleagues were familiar with existing guidelines, highlighting a critical gap in knowledge dissemination. This lack of clarity often led to inconsistent practice, reinforcing the need for clear, accessible policy communication and standardised education [22, 39]. Compounding this issue, the perceptions of perioperative team members regarding fasting often conflicted with actual clinical routines. In our study, this disconnect was evident in the varied interpretations and practices observed across surgical units. Similar findings were reported by De-Marchi et al., who noted that surgeons’ understanding of fasting recommendations did not align with actual patient experiences; for instance, the median recorded preoperative fluid fasting time was ten hours, with a range extending up to eighteen hours [38]. These prolonged fasting durations reflect a significant deviation from evidence-based recommendations and highlight a systemic issue in translating knowledge into practice.

Leadership gaps and rigid professional hierarchies further impeded the implementation of reduced fasting protocols. Both our findings and existing studies identified ambiguity over who holds primary responsibility for managing preoperative fasting as a recurring barrier [21, 35, 36]. This lack of clarity contributes to reluctance among staff to modify existing practices and perpetuates a culture of inaction [22, 36, 37].

Despite these challenges, the literature—and our data—emphasises several facilitators that support the successful adoption of reduced fasting interventions. These include targeted staff and patient education, improved communication across disciplines, and the demonstration of improved procedural efficiency because of protocol changes[40, 41]. Enhanced interdisciplinary collaboration and a shared understanding among clinicians were frequently cited as crucial enablers of change, reinforcing the importance of teamwork and open dialogue in driving sustainable practice improvements [22, 42].

### Strengths and limitations

This study recruited a mix of nursing and medical professionals, spanning anaesthetists, surgeons and geriatricians. However, it was conducted with primarily Australian clinicians and might not be generalisable to other countries and health systems internationally. We focussed on two topical programs aimed at reducing prolonged perioperative fasting practices (SipTilSend and OCL); however, there may be other programs that face different implementation barriers and enablers that were unable to be explored in our study.

## Conclusion

This study highlighted the required organisational and individual-level interventions for implementing evidence for preventing prolonged preoperative fasting in Australia. Our study pointed out the required steps for a successful implementation of the most appropriate and the most recent evidence to be translated into preoperative fasting practices. The main points focused on the governance and policy/procedure updates to support clinicians in adopting the change, addressing individual needs for knowledge dissemination and consideration and adapting the reduced fasting interventions to specific clinical contexts.

## Data Availability

The data are stored at the University of Sydney Research Data Management System and can be made available upon request.

## Acknowledgments

There are no competing interests to declare by any of the authors. Oya Gumuskaya was supported by the Australian College of Perioperative Nurses 2021 Research Grant, the Australian College of Perianaesthesia Nurses 2023 Research Grant. Oya Gumuskaya and Sarah Aitken were supported by the Sydney Health Partners Implementation Science Pilot Grant for this research. These data are stored at the University of Sydney Research Data Management System and can be available upon request. Mitchell Sarkies is supported by an NHMRC Investigator Grant (CIA Sarkies 2007970) and Sydney Horizon Fellowship. We acknowledge the collaboration and thank the SipTilSend network from Tayside, Scottland, UK, especially Dr Matthew Checketts, Sydney Health Partners Perioperative Care of Surgical Patients Clinical Academic Group, the Australian and New Zealand SipTilSend network, as well as all the clinicians from Sydney, Western Sydney, and Nepean Blue Mountains Local Health Districts of NSW, Australia.

